# Left subclavian artery revascularization with in-situ laser fenestration during TEVAR for complicated or high-risk TBAD: results of the LLTEVAR Trial

**DOI:** 10.1101/2025.02.20.25322187

**Authors:** Zhiyou Peng, Siyuan Liang, Rong Zhang, Peng Qiu, Minyi Yin, Guang Liu, Xiaobing Liu, Xinwu Lu, Kaichuang Ye

## Abstract

**Objective:** This study aimed to prospectively evaluate the safety and efficacy of left subclavian artery (LSA) revascularization via in-situ laser-assisted fenestration during thoracic endovascular aortic repair (TEVAR) for type B aortic dissection (TBAD).

**Methods:** This was a prospective, single-arm, multicenter study that enrolled patients with complicated or high-risk acute TBAD. The primary safety endpoint was freedom from composite major adverse events (mortality, stroke, myocardial infarction, paraplegia, type Ia endoleak) within 30 days after the procedure. Univariate and multivariate analyses determined the risk factors of composite major adverse events. The primary efficacy endpoint was freedom from all-cause death, LSA in-stent restenosis, and reintervention due to dissection progression, growth, and endoleak at 1 year after the procedure.

**Results:** A total of 100 patients (mean age, 60.5±12.6 years) were enrolled at 5 vascular centers between July 2018 and September 2021. The technical success rate of LSA revascularization was 98.0% (98/100). The primary safety endpoint within 30 days after the procedure was 86.0% (86/100, with 15 events occurring in 14 patients). Within 30 days after the procedure, mortality occurred in two patients, resulting in a 30-day mortality rate of 2.0% (2/100). Four patients experienced a stroke, and the rate was 4.0% (4/100). Retrograde aortic pathologies occurred in 2 patients, including one with ascending dissection treated with open repair, and the other with retrograde intramural hematoma treated with medical therapy, and the hematoma was self-resolved within 6 months. Five patients experienced type Ia endoleak, and conjunctive coil embolization were performed in 4 patients. One patient experienced paraplegia and recovered within 2 weeks. Complicated TBAD (OR, 4.98; 95%CI,1.27-19.55; P=.02) and type II/III arch (OR, 6.37; 95%CI,1.72-23.53; P=.006) significantly associated with a higher risk of composite major adverse events within 30-day after the procedure. The primary efficacy endpoint at 1 year after the procedure was 90.0%.

**Conclusion:** Left subclavian artery revascularization with in-situ laser-assisted fenestration during TEVAR for TBAD is a successful procedure and is associated with excellent short-term outcomes. However, attention should be paid to periprocedural major adverse events, especially in complicated TBAD patients with type II/III arch.

## INTRODUCTION

The procedure of thoracic endovascular aortic repair (TEVAR) has become the first-line intervention for patients with complicated and high-risk type B aortic dissection (TBAD) [1,2]. A healthy proximal landing zone has been confirmed to positively affect the short-term and long-term outcomes [3,4]. To achieve an adequate proximal landing zone in patients with a proximal entry tear originating in zone 2 and 3, coverage of the left subclavian artery (LSA) is required and may be associated with a higher incidence of stroke, spinal cord ischemia, vertebrobasilar ischemia and left arm ischemia [5–7]. Therefore, recent guideline recommended that LSA revascularization should be considered in selective patients during the TEVAR procedure in case of covering the LSA [8].

Current available methods for LSA revascularization during the TEVAR procedure include surgical bypass or transposition and endovascular techniques [9–11]. Surgical revascularization was once the standard approach; however, recent data show comparable periprocedural outcomes for endovascular and surgical techniques [8,12]. Furthermore, a significant high risk of complications of lymph leaks, nerve damage and other complications in patients with open bypass or transposition [13–16]. Endovascular techniques are less invasive approaches and are increasingly applied to reconstruct the LSA in conjunction with the TEVAR procedure. However, the parallel stent technique is associated with a high incidence of endoleak and stroke during the procedure [17], and the branched/fenestrated stent technique is still in the development phase and have some anatomical limitations with high cost [18]. In-situ laser-assisted fenestration for LSA revascularization might represent a commercially available and alternative endovascular technique.

The operative feasibility and technical details of in-situ laser-assisted revascularization for arch branches during TEVAR for aortic diseases in single center was retrospectively reported, which has become a widely used technique for the revascularization of LSA and other aortic branches in China [19–22]. We hypothesize that LSA revascularization using in-situ laser-assisted fenestration during TEVAR will not increase but rather decrease the incidence of major adverse events when compared to previously reported outcomes without LSA reconstruction. To investigate this, we conducted a prospective, single-arm, multicenter trial. The primary objective of this study was to assess the safety of in-situ laser-assisted fenestration for LSA revascularization during TEVAR in patients with acute TBAD involving a proximal entry tear originating in zones 2 and 3.

## METHODS

### TRIAL DESIGN AND PATIENTS

The LLTEVAR (LSA Revascularization With Laser-assisted Fenestration During TEVAR) study is a prospective, single-arm, multicenter trial that planned to enroll 100 patients at 5 vascular centers (NCT03845829). Patients presenting with complicated or high-risk acute TBAD with a proximal entry tear originating in zone 2 or zone 3, who were deemed appropriate candidates for TEVAR in conjunction with LSA revascularization, met the included criteria, and consented to participate were enrolled. The distance from the proximal edge of LSA orifice to the intended proximal landing location should be >5 mm. Patients were excluded if they were pre-procedurally scheduled to have proximal aortic arch coverage in zone 1 or zone 0. Other main exclusion criteria included patients who were inappropriate for TEVAR due to unfavorable anatomic characteristics (no commercially available aortic stent for the size of the aorta, no available access for TEVAR, both LAS and left vertebral artery originated from aorta, et al), previous open surgery for aortic arch and connective tissue disorders. The LLTEVAR trial was approved by the institutional review boards of Shanghai Ninth People’s Hospital, Shanghai Jiaotong University School of Medicine (SH9H-2018-T71-2), and all patients provided written informed consent. Reporting standards for TEVAR were based on the Society of Vascular Surgery guidelines [23].

### PATIENT MANAGEMENT

Once diagnosed, all patients received optimal medical therapy, consisting of close intensive care unit monitoring and control of blood pressure, heart rate, and pain. TEVAR procedures were performed for patients with complicated and high risk TBAD. Patients not responding to medical therapy or presenting with rupture or impending rupture, malperfusion were treated emergently; otherwise, TEVAR procedures were performed 2 weeks later. The procedure of TEVAR in adjunct with in-situ laser-assisted fenestration for LSA revascularization has been previously described in detail [19–21]. Briefly, the TEVAR procedures were performed under local or general anesthesia, and aortic stent-grafts were deployed through femoral access, and the LSA fenestration stents were inserted through brachial artery. The diameter of the aortic stent-graft was based on the diameter of the proximal healthy landing zone of the aorta with approximately 5% oversizing. If the diameter of the stent-graft mismatched the diameter of the distal attachment site of the aorta (e.g, > 20% oversizing of the total aortic diameter), a distal restricted stent technique was used [24]. After aortic stent-graft deployed, a 6F steerable sheath combined with a balloon catheter (Mustang, Boston Scientific, 4mm in diameter and 4cm in length) allowed the laser tip to be placed in the intended fenestration site outside of the aortic stent-graft. After the fenestration was established, the balloon catheter was pushed forward and through the fenestration hole, and after the balloon angioplasty for the established hole, the laser tip was exchanged with a 0.035-inch guidewire. Then the fenestration hole was subsequently dilated up to the reference diameter of the subclavian artery, and a balloon-expandable (Lifestream, BD) or self-expandable (Fluency, BD) covered-stent (10% oversizing of the LSA in diameter) was deployed inside the fenestration. Post stent dilation was always required for the LSA stent. At the end of the procedure, arteriography was performed from the ascending aorta to assess the success of the procedure and to identify potential endoleak and residual stenosis of the LSA stent.

All patients were discharged with oral medications to maintain blood pressure and heart rate in a stable condition. Aortic CT angiography was routinely performed before discharge, at 1 month, at 6 months, at 12 months and then annually during the extended follow-up. A follow-up CT angiography was also performed when patients experienced symptoms recurrence and suspicious symptoms associated with aortic pathologies.

### TRIAL ENDPOINTS

The primary safety endpoint was freedom from composite major adverse events (MAEs) within 30 days after the procedure. MAEs included mortality, paraplegia, stroke, myocardial infarction, type Ia endoleak, and retrograde dissection/IMH. A safety committee adjudicated the endpoints in this trial. Previous reports indicated that the composite major adverse event rate within 30 days for patients undergoing TEVAR for TBAD without LSA revascularizations was 25% [25]. We hypothesized that incorporating LSA revascularization could reduce the composite major adverse event rate to 15%. Using an 80% power and a one-sample log-rank test with a 10% one-sided alpha for sample size calculation, we determined that 80 patients would be necessary for safety evaluation. To account for an anticipated 20% loss to follow-up, we planned to recruit a total of 100 patients. Univariate and multivariate analyses were used to identify the potential risk factors (general characteristics of patients, anatomic information of dissection, stage of TEVAR procedure) for MAEs within 30 days after the procedure. The primary efficacy endpoint was freedom from all-cause death, LSA in-stent restenosis, and reintervention due to dissection progression, growth and endoleak at 1 year after the procedure.

## STATISTICAL ANALYSIS

Continuous variables are presented as mean with standard deviation, and categorical variables are summarized using counts and proportions. Univariate logistics regression analysis was performed to determine the potential risk factors for MAEs within 30 days after the procedure, and only factors found statistically significant in the univariate analysis were included in the multivariate analysis. The primary efficacy endpoint at 12 months after the TEVAR procedure was estimated using the Kaplan-Meier method. SPSS version 26.0 (SPSS, Inc., Chicago, IL, USA) was used for the statistical analyses. Differences with a P value <.05 were considered statistically significant.

## RESULTS

### BASELINE CHARACTERISTICS AND PROCEDURAL CHARACTERISTICS

Between July 2018 and September 2021, a total of 100 patients were enrolled at 5 vascular centers. Patients’ general characteristics and comorbidities are detailed in Table 1. More than two-thirds of patients were treated under local anesthesia. Aortic stent-graft implantation was performed through femoral artery puncture, except for one patient who underwent incision. Technical success of the TEVAR procedure was achieved in all patients, with correct deployment of the aortic stent-grafts and complete exclusion of the proximal entry tear. However, intentional coverage of the left carotid arteries during TEVAR procedure in order to achieve an adequate proximal landing zone was required in 4 patients, who were simultaneously treated with parallel stenting, and no major adverse events were observed in these 4 patients. The technical success of in-situ laser-assisted fenestration for LSA revascularization was 98.0% (98/100), with parallel stenting performed in the remaining 2 patients with failed laser-assisted fenestration due to inadequate distance from the proximal edge of the aortic graft to the intended fenestration site (Table 2). Three superior mesenteric arteries in 3 patients presenting with mesenteric ischemia were reconstructed with bare mental stents. Six patients had common iliac artery revascularization due to limb ischemia before the TEVAR procedure. Distal restricted stents were required in 12 patients. Coil embolization was simultaneously undertaken in 4 patients with type Ia endoleaks after TEVAR (Video 1-4). Prophylactic cerebrospinal fluid drainage was performed pre-procedurally in one patient who had prior EVAR for abdominal aorta pathology. Nevertheless, the patient experienced paraplegia on the first day after TEVAR and achieved full recovery before discharge. The specifics of the procedure were presented in Table 2.

**TABLE 1.**
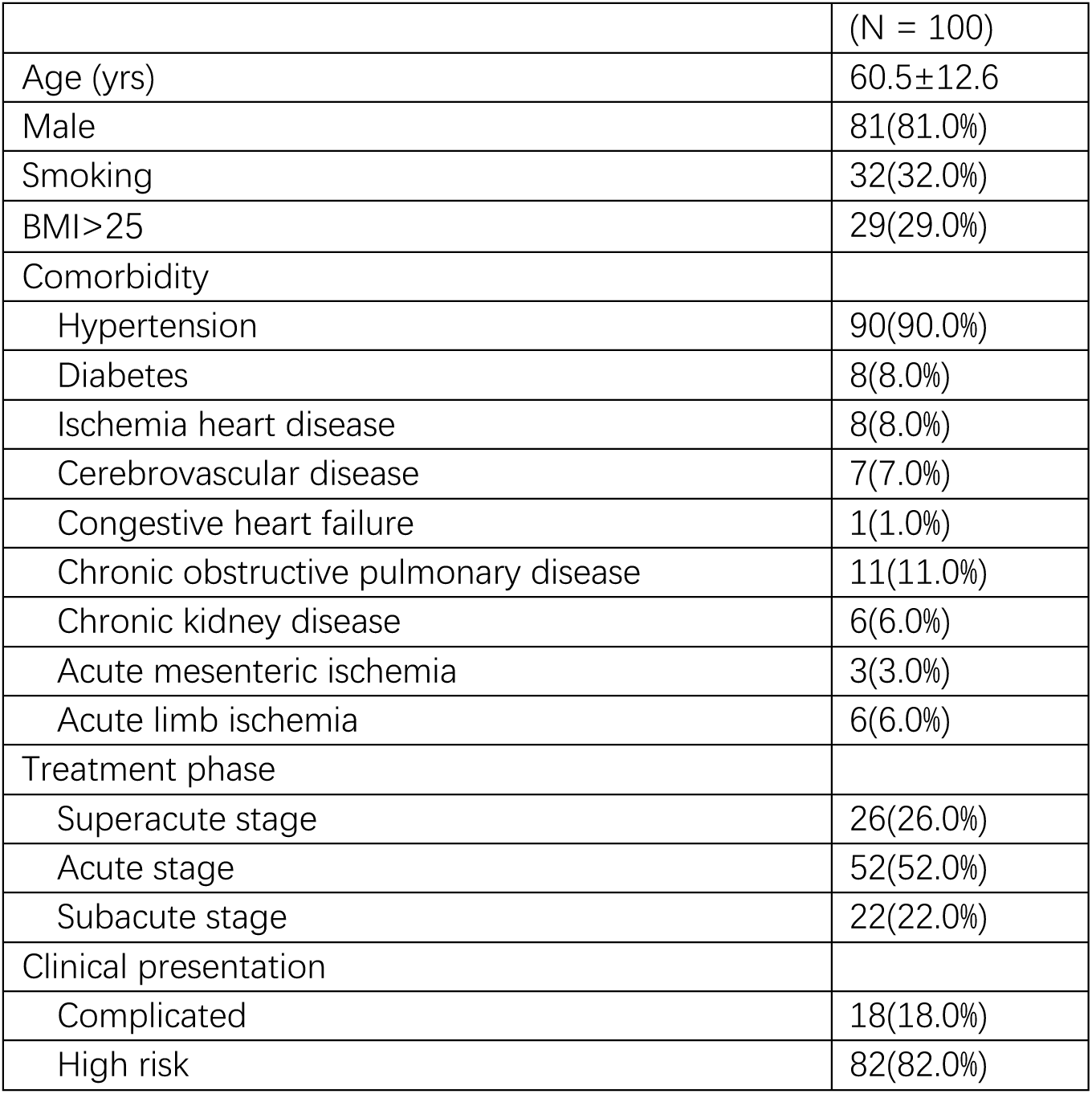
Baseline characteristics before procedures.

**TABLE 2.** Procedural Details of TEVAR in conjunctive with LSA reconstruction.

|  |  |
| --- | --- |
| Procedural details | N = 100 patients |
| Aortic stent-grafts |  |
| GORE TAG | 43(43.0%) |
| MEDTRONIC VALIANT | 19(19.0%) |
| Other stent-grafts | 38(38.0%) |
| LSA fenestration stent |  |
| GORE Viabahn | 1(1.0%) |
| BD Lifestream | 13(13.0%) |
| BD Fluency | 86(86.0%) |
| Adjunctive procedures |  |
| Parallel stent for left subclavian artery | 2(2.0%) |
| Parallel stent for left carotid artery | 4(4.0%) |
| Visceral artery reconstruction with stent | 3(3.0%) |
| Common iliac artery reconstruction with stent | 6(6.0%) |
| Preventive cerebrospinal fluid drainage | 1(1.0%) |
| Distal tapered restrictive covered stent | 12(12.0%) |
| Embolization with coils | 4(4.0%) |

### SAFETY OUTCOMES

The primary safety endpoint was 86.0% (86/100), with 15 events occurring in 14 patients (Table 3). Type Ia endoleak was identified in five patients. Of these, four received simultaneous treatment with coil embolization, while the remaining patient underwent close surveillance. Four patients experienced a stroke, and one of them died 6 days after the procedure. One patient died 4 days later after the procedure due to mesenteric ischemia leading to the 30-day mortality rate of 2.0% (2/100). One patient experienced paraplegia. The patient was treated with CSF drainage, and recovered within 2 weeks. One patient presented with myocardial infarction 2 days after the TEVAR procedure and was treated with coronary artery stenting emergently. Retrograde ascending aortic dissection occurred in one patient, and the patient was treated with surgical repair. One patient presented with retrograde ascending aortic intramural hematoma and was treated with medical therapy, and the intramural hematoma partially and completely resolved in 1 month and 6 months, respectively (Figure 1).

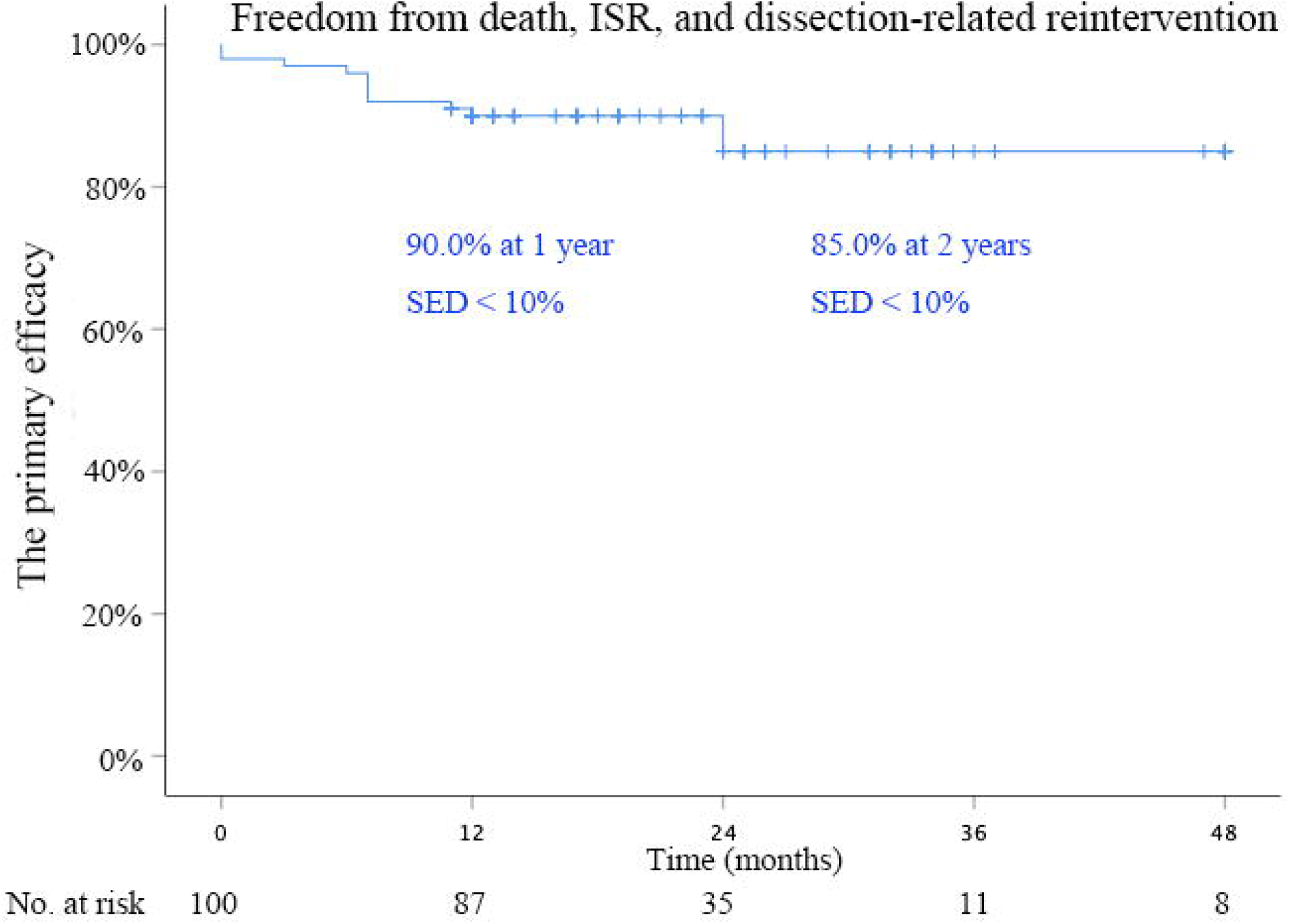

**TABLE 3.** Major adverse events within 30 days after procedure.

|  |  |
| --- | --- |
| Major adverse events | 15 events occurred in 14 patients |
| Death | 2(2.0%) |
| Stroke<sup>1</sup> | 4(4.0%) |
| Myocardial infarction | 1(1.0%) |
| Ia endoleak<sup>3</sup> | 5(5.0%) |
| Spinal cord ischemia<sup>4</sup> | 1(1.0%) |
| Retrograde dissection<sup>5</sup> | 1(1.0%) |
| Retrograde IMH<sup>6</sup> | 1(1.0%) |

1:one patient presented with stroke died within 1 week after procedure;
2:one patient presented with mesenteric ischemia died 4 day after procedure;
3:Type Ia endoleak occurred in 5 patients and 4 of them were treated with coil embolization.
The remaining one presented with type Ia endoleak during the follow up, and was successfully treated with coil embolization at 7 months after TEVAR procedure;
4. one patient presented with paraplegia and was successfully treated with CSF drainage;
5. this patient was successfully treated with surgical repair;
6. this patient was successfully treated with medical therapy.

### Multivariable analysis for MAEs

Univariate logistic regression was performed to identify the potential risk factors for composite MAEs within 30 days after the procedure (Table 4). Complicated TBAD (OR, 3.60; 95%CI,1.10-11.74; P=.03), LSA-aorta angle <45 (OR, 4.33; 95%CI,1.07-17.64; P=.04) and arch type II/III (OR, 6.00; 95%CI,1.89-19.04; P=.002) were enrolled for further multivariate logistic regression. The result of multivariate logistic regression identified complicated TBAD (OR, 4.98; 95%CI,1.27-19.55; P=.02) and arch type II/III (OR, 6.37; 95%CI,1.72-23.53; P=.006) as two independent risk factors for MAEs within 30 days after the procedure.

**TABLE 4.** Univariate and multivariate analysis of potential risk factors for major adverse events.

| Risk factors | Univariate |  |  |  | Multivariate |  |
| --- | --- | --- | --- | --- | --- | --- |
|  | without MAE (86) | with MAE (14) | OR (95% CI) | P value | OR (95% CI) | P value |
| General characteristics |  |  |  |  |  |  |
| Age | 60.5±12.1 | 60.4±13.9 | 1.00(0.96-1.04) | 0.98 |  |  |
| Gender (male) | 67 (79.8%) | 14 (87.5%) | 1.78(0.37-8.57) | 0.48 |  |  |
| Hypertension | 74 (88.1%) | 15 (93.8%) | 2.03(0.24-17.04) | 0.52 |  |  |
| Smoking | 31 (36.9%) | 7 (43.8%) | 1.33(0.45-3.93) | 0.61 |  |  |
| BMI>25 | 24 (28.6%) | 5 (31.3%) | 1.14(0.36-3.62) | 0.83 |  |  |
| Complicated* | 12 (14.3%) | 6 (37.5%) | 3.60(1.10-11.74) | 0.03 | 4.98(1.27-19.55) | 0.02 |
| Dissection anatomic |  |  |  |  |  |  |
| Tear located in inner aortic curve | 16 (19.0%) | 4 (25.0%) | 1.42(0.40-4.97) | 0.59 |  |  |
| Diameter of proximal tear >10 mm | 26 (30.95%) | 8 (50.0%) | 2.23(0.76-6.59) | 0.15 |  |  |
| FL diameter > 22 mm | 32 (38.1%) | 6 (37.5%) | 0.98(0.32-2.94) | 0.96 |  |  |
| Distance between LSA and proximal tear < 2cm | 37 (44.0%) | 10 (62.5%) | 2.12(0.71-6.36) | 0.18 |  |  |
| Proximal entry tear in zone 2 | 6 (7.1%) | 1 (6.25%) | 0.87(0.10-7.73) | 0.90 |  |  |
| LSA-Aorta angle <45 | 6 (7.1%) | 4 (25.0%) | 4.33(1.07-17.64) | 0.04 | 2.69(0.49-14.75) | 0.26 |
| Arch type II/III* | 12 (14.3%) | 8 (50.0%) | 6.00(1.89-19.04) | 0.002 | 6.37(1.72-23.53) | 0.006 |
| Procedure characteristics |  |  |  |  |  |  |
| Time of intervention (<14 days) | 66 (78.6%) | 12 (75.0%) | 0.82(0.24-2.84) | 0.75 |  |  |
| Type of stent-graft (GORE) | 37 (44.1%) | 6 (37.5%) | 0.76(0.25-2.29) | 0.63 |  |  |
| Type of LSA stent (Fluency) | 72 (85.7%) | 14 (87.5%) | 1.17(0.24-5.79) | 0.85 |  |  |
| Distal restrict stent | 10 (11.9%) | 2 (12.5%) | 1.06(0.21-5.35) | 0.95 |  |  |

\*The risk of periprocedural major adverse events of a complicated TBAD patients presented with type II/III arch is 7.96 (95% CI 2.33-27.19; P=0.001).
BMI: Body Mass Index. CI: Confidence interval. FL: False lumen. LSA: Left subclavian artery.
OR: odds ratio.

### EFFICACY OUTCOMES

The primary efficacy endpoint at 1 year after procedure was 90.0% (Figure 2), and there was no significant difference in the primary efficacy between patients with and without periprocedural major adverse events (91.6% vs. 81.3%, P=.39. Figure 3). After a mean follow-up of 20.4 months (range, 1-60 months), no stent-graft migration and stent fracture were identified in CT angiography. Two patients died during the follow up, including one patient who died of brain hemorrhage at 5 months and the other patient who died of acute coronary syndrome at 7 months after the procedure. Two patients experienced LSA in-stent restenosis at 6 months and 7 months after the procedure, respectively, and one of them was successfully treated with balloon angioplasty. Two patients who complained of chest pain underwent coil embolization, one for a type Ia endoleak (at 7 months) and the other for a type II endoleak (at 11 months). Type R endoleak occurred in 4 patients, and three patients of them were treated with additional distal aortic stent-graft (at 12, 24 and 24 months after the TEVAR procedure, respectively), while the remaining one was managed with closely surveillance due to the absence of symptoms and false lumen expansion during the follow-up. One patient required a FEVAR at 3 months after the TEVAR procedure due to the expansion of the abdominal aorta.

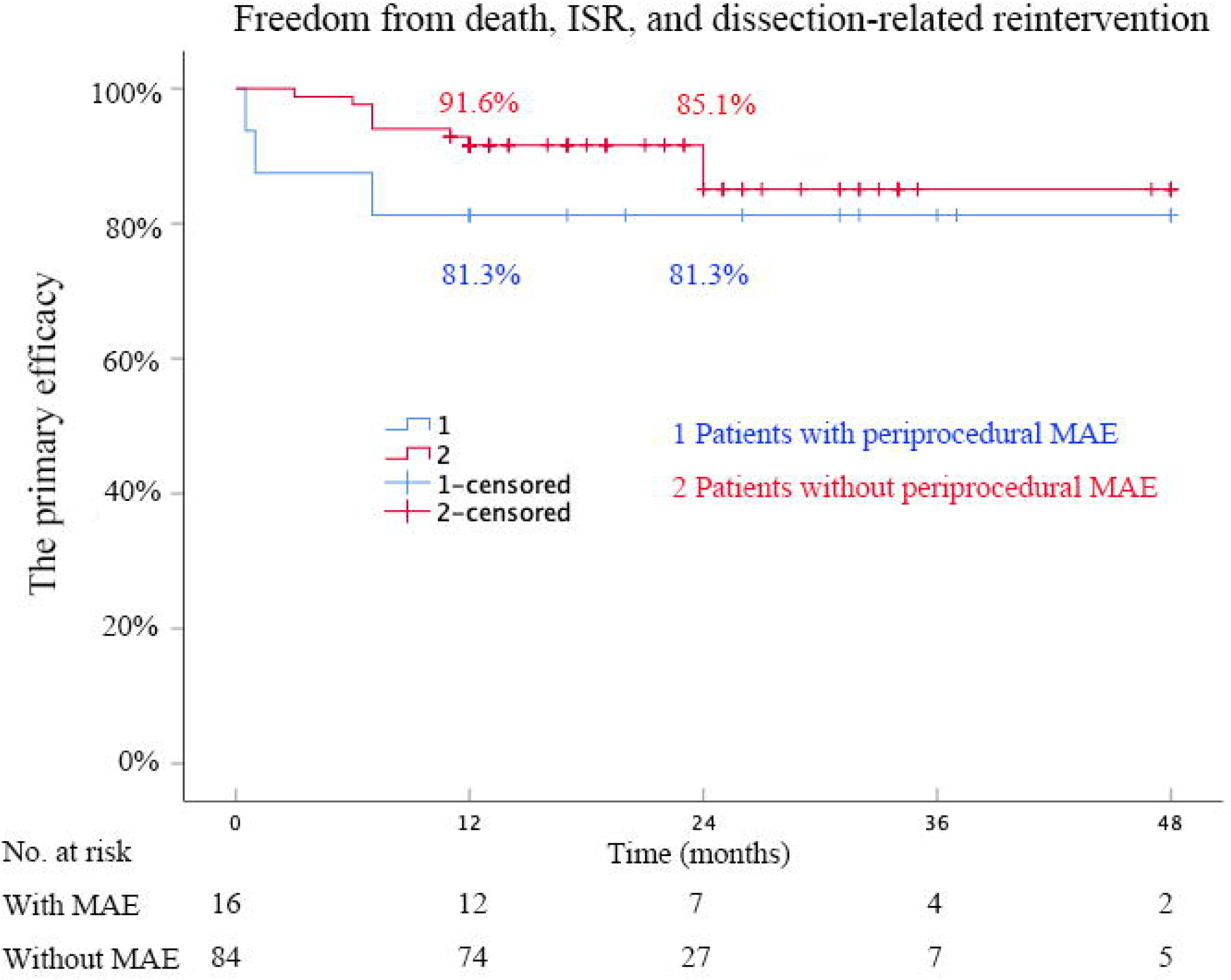

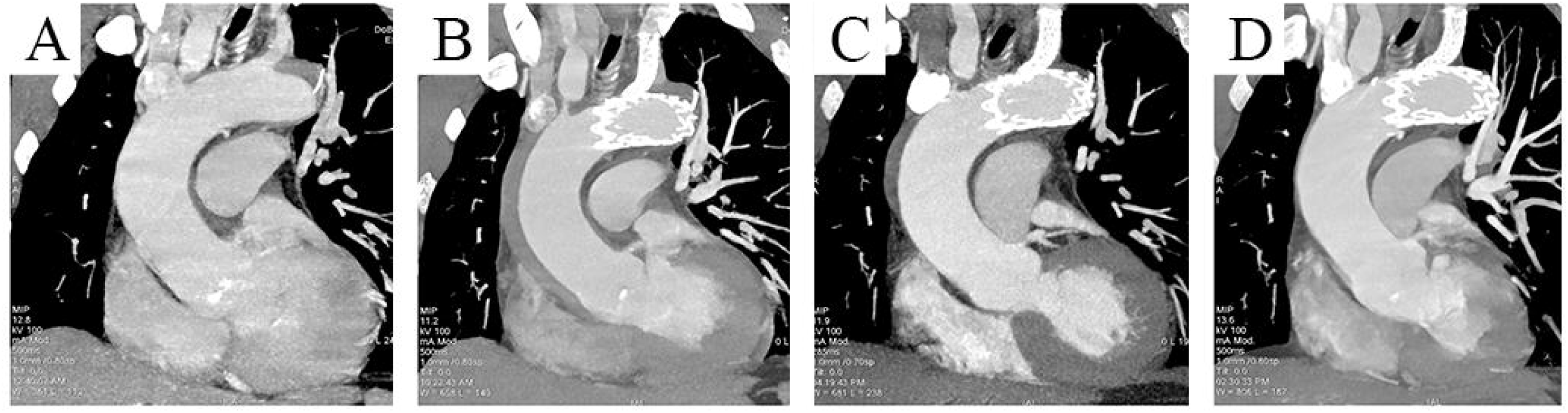

## DISCUSSION

The outcomes of this prospective study demonstrated that in-situ laser-assisted fenestration for LSA revascularization in conjunctive with the TEVAR procedure for complicated and high risk acute TBAD is a highly successful procedure associated with excellent short-term outcomes. However, periprocedural MAEs, especially immediate type Ia endoleak and stroke after the procedure in complicated TBAD patients with type II/III aortic arch, cannot be ignored. Fortunately, type Ia endoleak could be successfully treated with simultaneous coil embolization.

In patients with a short distance of the proximal intimal tear from the LSA, especially in patients with a proximal tear in zone 2 and zone 3, the coverage of the LSA is warranted to achieve an adequate proximal landing zone [3–7]. However, the TVEAR procedure with LSA coverage was associated with a twofold higher risk of major adverse events compared with TVEAR procedure without LSA coverage [26], and the most frequent adverse events was type II endoleak and persistent flow within the false lumen [27]. Although, embolization of LSA could decline the incidence of endoleak, the consequences were obvious, such as a trending increase in the risk of paraplegia (OR, 2.69; 95%CI, 0.75-9.68) and anterior circulation stroke (OR, 2.58; 95% CI, 0.82-8.09), and a significant increase in the risk of arm ischemia (OR, 47.7; 95% CI, 9.9-229.3) and vertebrobasilar ischemia (OR, 10.8; 95% CI, 3.17-36.7) [25]. Furthermore, some TBAD patients need to undergo EVAR procedure in the chronic stage, and it is obvious that the risk of paraplegia is further increased. Hence, current guidelines recommend revascularization of the LSA if TEVAR coverage obstructed antegrade LSA flow [8,28].

Type Ia endoleak emerged as the most prevalent MAEs in this study. Of the 14 patients who encountered major adverse events, 5 experienced type Ia endoleak, resulting in an overall incidence rate of 5%. The occurrence of Type Ia endoleak exhibited notable variations across different procedures. According to a meta-analysis [17], individuals undergoing TEVAR with chimney exhibited the highest risk of type Ia endoleak at 18.8% (182 occurrences out of 964 patients), while those undergoing TEVAR with in-situ fenestration had the lowest rate at 2.2% (12 out of 528 patients). Our findings indicate a relatively elevated incidence of this complication, possibly attributable to a higher number of patients with type II/III aortic arch. Piazza M et al. reported that a significantly greater proximal sealing length may be necessary to reduce the risk of type Ia endoleak during TEVAR in patients with type II/III aortic arches [4]. Fortunately, 4 patients with type Ia endoleak were found during the TEVAR procedure and were successfully treated with coil embolization. The remaining untreated one case of type Ia endoleak still existed during follow-up and was also successfully treated with coil embolization at 7 months after TEVAR procedure.

Stroke is another periprocedural MAEs in this study. Although all LSAs were reconstructed in the TEVAR procedure, the incidence of stroke was still as high as 4.0%, which aligns with previously reported estimates, ranging from 2.7% to 5.8% in meta-analyses and larger registries [7,29,30]. Besides the coverage of LSA without revascularization, which will increase the risk of stroke, patients with complex arch anatomy (type II/III arch) are more susceptible to stroke events after TEVAR [31]. Meanwhile, one meta-analysis revealed that the incidence of stroke after TEVAR procedure in complicated TBAD was higher than in uncomplicated TBAD (5.85% vs. 3.92%; P<.01) [30]. Although uncomplicated TBAD patients were not included in the current study, the incidence of MAEs was higher in complicated TBAD than that in high-risk TBAD. One of the reasons is that the TEVAR procedure for complicated TBAD was always performed in superacute or acute stages, which might increase the incidence of MAEs. Li et al found that patients treated in acute had a significantly higher risk of MAEs within 30 days after TEVAR, with 15.8% in acute compared to patients with 5.2% in the subacute (P=.03) [32]. In one meta-analysis [33], patients with TBAD treated in hyperacute encounter a higher risk of MAEs within 30 days after the procedure, with an odds ratio of 1.58 (95% CI 1.12-2.24, P=.009) compared with late interventions.

In patients with type II/III aortic arch, a longer proximal seal length is required for a safe and durable TEVAR procedure for acute TBAD [4]. In the current study, the aortic stent graft covered the LSA in all patients to extend the proximal seal length. Despite this, sixteen MAEs still occurred in 14 patients within the initial 30 days post-procedurally, and individuals with complicated TBAD and type II/III aortic arch experienced a higher incidence of MAEs. This phenomenon is multifactorial. Firstly, it is associated with the heightened technical complexity of performing in-situ fenestration for LSA revascularization, which inevitably extends the overall duration of the procedure. The iterative maneuvers within the arch during procedure evidently contribute to an increased risk of stroke. Secondly, these findings underscore the imperative for innovative approaches and modifications in the design of aortic stent grafts tailored to patients with type II/III aortic arch, taking into careful consideration the complexity identified in this subset of individuals.

### LIMITATIONS

This study has several limitations. First, a major limitation of the present study was the absence of a comparator group. Consequently, the safety and efficacy results were not compared with carotid-subclavian bypass and transposition, which were traditional standard approaches. However, in the current endovascular era, preserving the normal anatomical structure of LSA is crucial for potential access site in the fenestrated and branched EVAR procedure. Second, this study investigated a short follow-up period, and some negative efficacy outcomes may occur with a longer follow-up. It has been reported that approximately half of the dissection will progress to aneurysmal dilation of the dissected aorta at 5 years after the initial onset of dissection, and one-third of patients require intervention for aneurysmal pathologies of the diseased aorta [34,35]. Therefore, a long-term follow-up is necessary to reveal the extended results after the TEVAR procedure. Third, while several risk factors for MAEs after the procedure were identified, hemodynamic variables were not assessed in this study, requiring further investigation in future studies.

## CONCLUSIONS

In this prospective, single-arm, and multicenter trial, the in-situ laser-assisted fenestration of LSA revascularization in conjunctive with the TEVAR procedure for complicated and high risk acute TBAD with a proximal entry tear originating in zone 2 and 3 is a successful procedure associated with excellent short-term outcomes. Attention should be directed towards patients with type II/III aortic arch and those with complicated TBAD, given the higher risk of periprocedural stroke and type Ia endoleak. Further studies incorporating comparator groups and longer-term follow-up are warranted to validate these promising results. Additionally, forthcoming aortic stent-grafts should be designed to mitigate the risk of MAEs within 30 days after TEVAR for complicated TBAD with complex arch anatomy.

### Contributions

We sincerely thank Doctor Wei Xiong (Wuhan No. 1 Hospital, Hubei, China), Doctor Zhijun He (Zhoupu Hospital, Shanghai, China) and Qun Huang (Shanghai Ninth People’s Hospital, Shanghai, China) for the warm help they gave to this trial.

Disclosures: The authors declared they do not have anything to disclose regarding the conflict of interest for this manuscript.

Funding: This study was financially supported by the National Natural Science foundation of China (Grant No. 82370494), and the Science and Technology Committee of Shanghai (Grant No. 21S31904300), and the Clinical Research Program of Shanghai Ninth People’s Hospital, Shanghai JiaoTong University School of Medicine (Grant No. YBKB202224).

## Data Availability

All data produced in the present study are available upon reasonable request to the authors.

